# Imaging Infants and Children with Investigational Handheld Optical Coherence Tomography with Widefield Lens: A Pilot Study

**DOI:** 10.1101/2024.10.30.24316132

**Authors:** Angela S. Li, Ryan Imperio, Du Tran-Viet, Shwetha Mangalesh, S. Grace Prakalapakorn, Lejla Vajzovic, Ramiro Maldonado, Sharon F. Freedman, Xi Chen

## Abstract

Handheld optical coherence tomography (OCT) systems have shown promises to provide detailed evaluation of the pediatric retina. It is less stressful for preterm infants compared to binocular indirect ophthalmoscopy and shows promises for screening for retinopathy of prematurity. There are currently only one commercially available non-contact handheld OCT systems, and no commercially available widefield handheld OCT systems. Here we report and evaluate the first use of the Theia Imaging Investigational handheld OCT system with widefield lens (T1-W, Theia Imaging, Durham, NC) in pediatric subjects. We compare the OCT en face view to handheld widefield or tabletop ultra-widefield fundus photographs and evaluate the ability to visualize the vitreoretinal interface, intraretinal, subretinal and choroidal features on cross-sectional OCT scans.

## Introduction

Optical coherence tomography (OCT) imaging has become an important imaging modality to evaluate the pediatric retina. It is a standard-of-care instrument to evaluate and monitor macular pathology and thicknesses in children who are able to cooperate with tabletop imaging in clinic, and has increased our understanding of the macular pathology of predominantly peripheral retinal diseases.^1^ Handheld OCT has allowed for imaging of infants and young children in a supine position and during anesthesia. Usage of handheld OCT was first described in infants with shaken baby syndrome^2^ and at bedside in preterm infants with retinopathy of prematurity (ROP).^3,4^ Imaging of infants with handheld OCT has shed light on macular edema in the preterm infants,^5-7^ and retinal morphology at the vascular-avascular junction in the peripheral retina in infants with ROP.^8-11^ Handheld OCT also leads to less infant stress compared to binocular indirect ophthalmoscopy as it does not emit a bright light.^12^

Currently, there is only one commercially available handheld OCT device (Bioptigen/Leica Envisu C2300) which was approved by the FDA more than 10 years ago. An armature-fixated non-contact OCT device recently received FDA clearance in the US (Heidelberg FLEX, Heidelberg, Germany). However, the utility of both systems is limited by the field-of-view (FOV, 40 degrees), the size and weight of the probe/system, and the long image capture time. Other handheld OCT imaging systems are currently in investigational use.^13-17^ Many non-contact systems have a limited FOV, especially compared to the contact fundus imaging (RetCam photography), which is often used in aid to or replace a clinical exam with indirect ophthalmoscopy. The objective of this pilot study is to evaluate the utility of a new, contact widefield swept-source OCT system (Theia Imaging, LLC, Durham, NC) with widefield lens in capturing en face and cross-sectional OCT images in pediatric patients undergoing exam under anesthesia and infants in the clinic or the nursery.

## Results

We captured widefield swept-source OCT in 26 eyes of 14 pediatric subjects (age range 36 weeks post menstrual age preterm to 15 years-of-age) using the Theia T1-W investigational handheld OCT system with widefield lens. 24 eyes of 13 patients were imaged during examination under anesthesia (EUA) and 4 eyes of 2 patients were imaged at bedside in the NICU or in clinic (2 eyes of 1 subject had two imaging sessions: once in clinic and once during EUA).

### Widefield retina en face visualization

In all eyes, we were able to capture the fovea, optic nerve, and mid-peripheral retina in a single OCT volume including awake infants at the bedside. The FOV of the OCT en face image captured with investigational handheld OCT system with widefield lens was of good quality and was larger than the widefield fundus photograph (RetCam, Natus, Middleton, WI) **(Figure 1A-C)**. Of note, the vessel visualization toward the periphery of the en face image remains clear, while in the fundus photograph, the view toward the periphery of the image was darker. Tilting the lens allowed more peripheral view to the retina to equator compared to the tabletop ultra-widefield fundus photography (Optos, Dunfermline, Scotland) **(Figure 1D-F)**.

**Figure 1.**
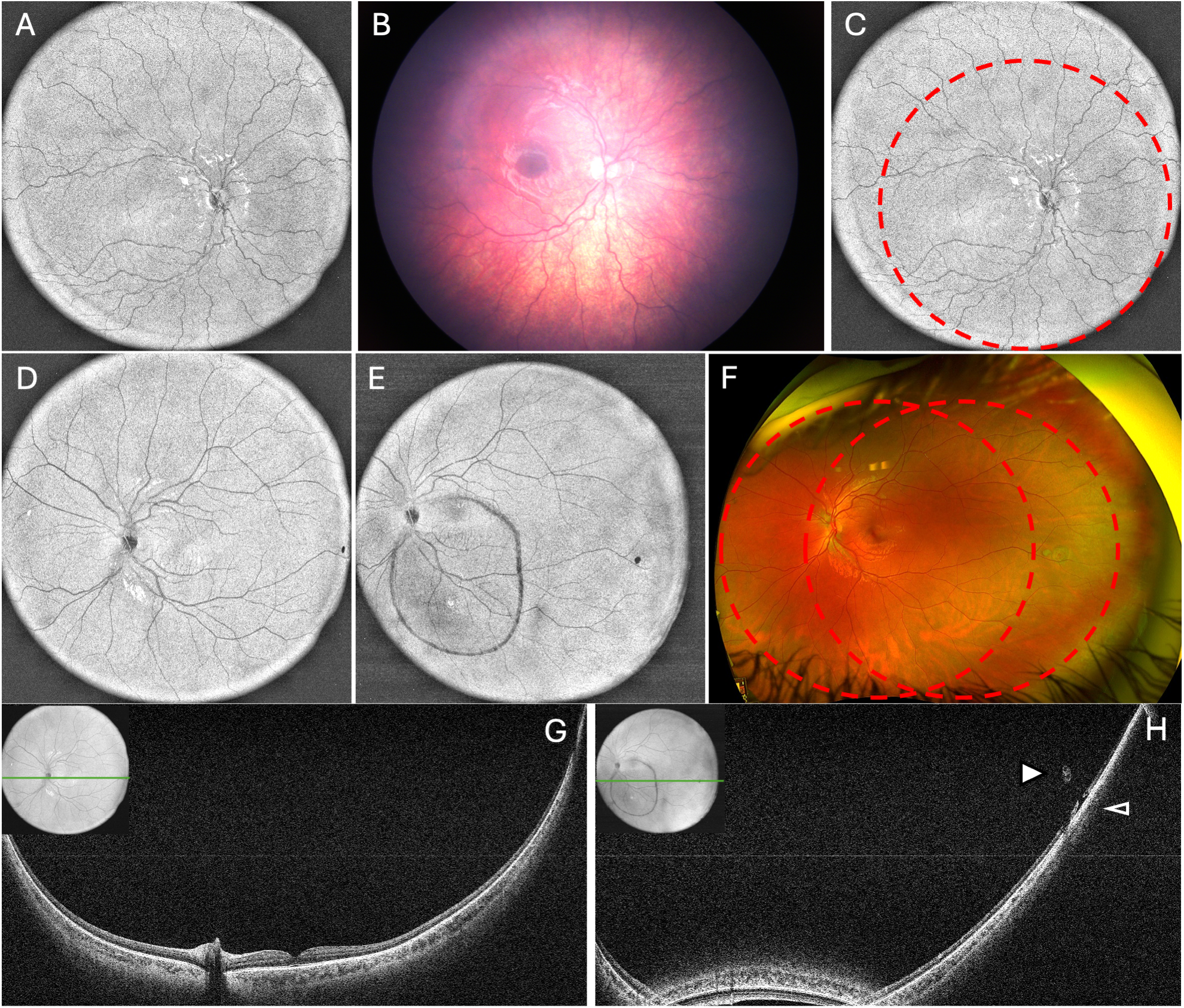
Comparison of the field-of-view of widefield en face OCT with investigational handheld OCT system and handheld widefield and tabletop ultra-widefield fundus photography. A-C. Right eye of a young child with optic nerve hypoplasia was imaged using widefield OCT (A and C) and widefield fundus photography (B). For comparison, the corresponding field-of-view of the fundus photography was outlined on the en face OCT capture (red dashed line, C). D-H. Left eye of a child with temporal operculated hole was imaged with widefield en face OCT (D, E), cross-sectional OCT (G, H) and tabletop ultra-widefield fundus photography (F). For comparison, the corresponding field-of-view of the en face OCT was outlined on the ultra-widefield fundus photograph (red dashed line, F). Cross-sectional OCT scan through the fovea (G) and the temporal operculated hole (H) visualized focal subretinal fluid at the operculated hole (open arrowhead) and the overlying operculum (closed arrowhead). The location of the cross-sectional OCT scan was marked with green line on the en face scan inset.

### Widefield cross-sectional visualization of the vitreoretinal interface

The investigational handheld OCT system with widefield lens allowed unique visualization of the vitreoretinal interface that was not well seen on clinical exam, fundus photography, fluorescein angiography or ultrasound. Vitreoretinal features visualized on wide-field cross-sectional OCT scans includes the vitreous traction at the operculated hole **(Figure 1H)**, vitreous traction over focal retinal thickening superior to the optic disc in a child with Coats’ disease **(Figure 2D)**, over localized focal retinoschisis and over areas of retinal detachment **(Figure 3D)** in a child with history of ROP presenting with retinal detachment **(Figure 3C)**, multifocal vitreous traction over the posterior pole in infants status post retinal detachment repair **(Figure 4C, D and Figure 5B-E)**, extraretinal neovascularization in stage 3 ROP and its associated vitreous traction **(Figure 6E-H and supplementary information)**, and vitreous hyperreflective foci **(Figure 2D,E and Figure 6E-H)**.

**Figure 2.**
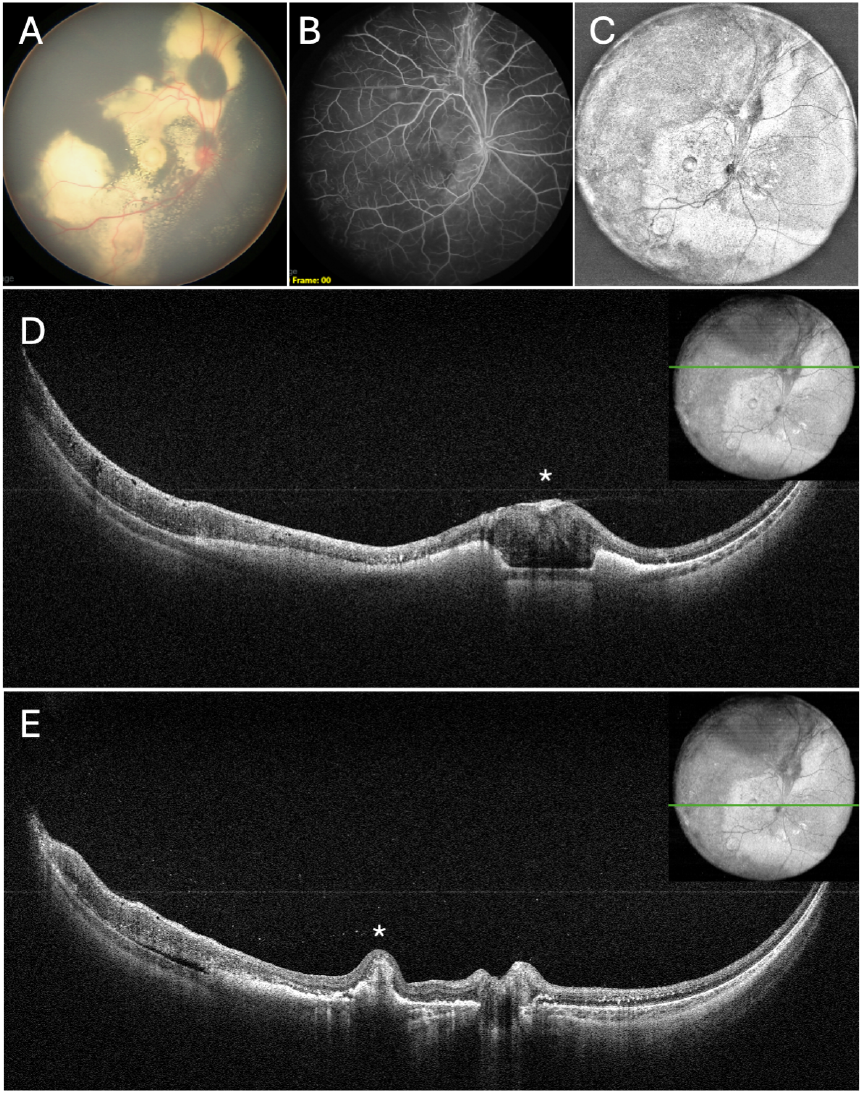
Right eye of a young child with Coats’ disease imaged under anesthesia. Widefield fundus photograph (A) showed extensive exudates in the posterior pole, with a central macular nodule and a nodule above the optic disc. Fluorescein angiography (B) demonstrated vascular contraction around the superior nodule. En face OCT (C) visualized the nodule as well as shaded area temporally consistent with retinal thickening on cross-sectional OCT scans. Cross-sectional OCT scan through the superior nodule (D, asterisk) showed localized epiretinal membrane and associated vitreous traction, vitreous hyperreflective foci, intraretinal and subretinal hyperreflective material, and temporal retinal thickening. Cross-sectional OCT scan through the macula (E) showed central nodule associated with RPE and hyperreflective elevation (asterisk), vitreous hyperreflective foci, intraretinal and subretinal hyperreflective material, temporal retinal thickening and thin layer of subretinal fluid. The location of the cross-sectional OCT scan was marked with green line on the en face scan inset.

**Figure 3.**
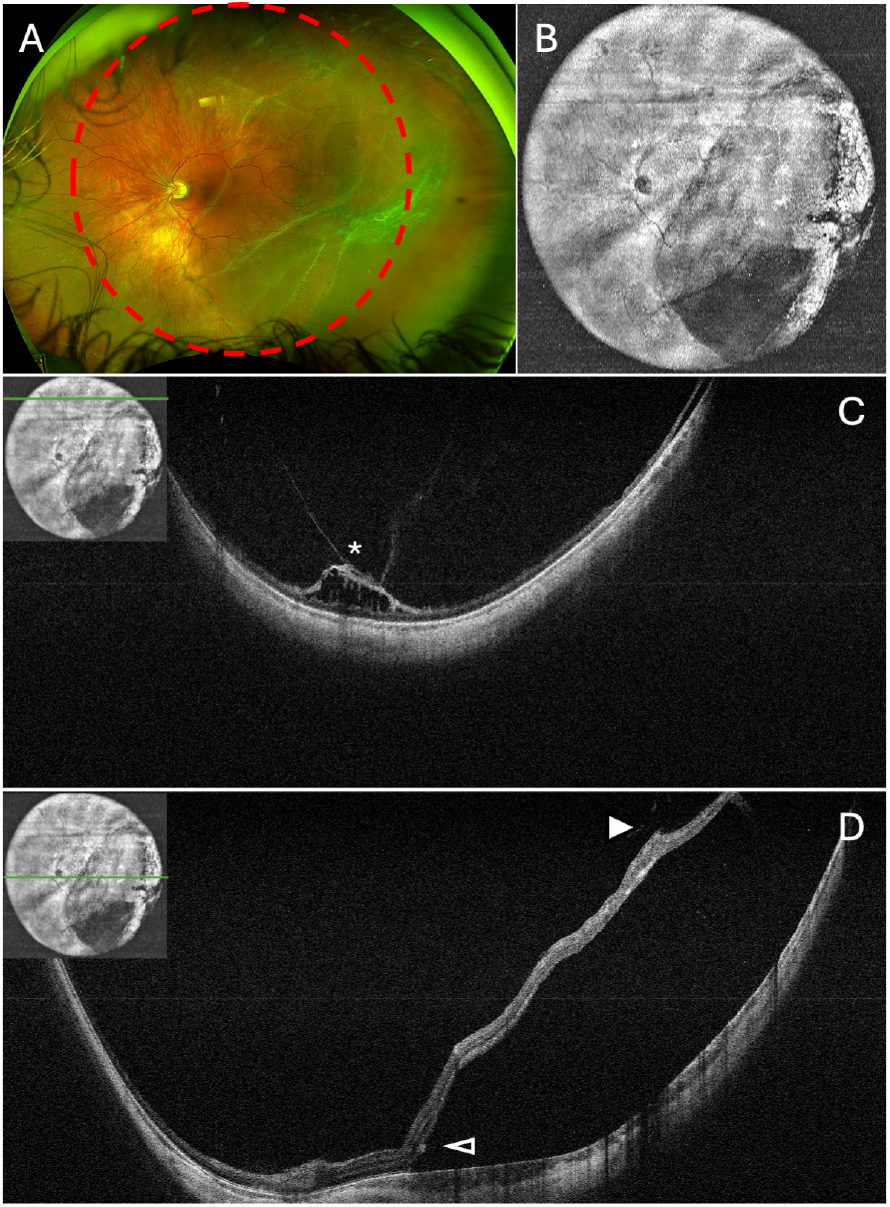
Left eye of a child with history of untreated retinopathy of prematurity and temporal retinal detachment imaged under anesthesia. The OCT en face view (B) showed temporal shaded area consistent with area of retinal detachment, and its corresponding location was marked on the ultra-widefield fundus photograph (A). Cross-sectional OCT scan through the superior retina (C) showed an area of localized tractional retinoschisis (asterisk) not appreciated on clinical exam. Cross-sectional OCT scan through the macula showed temporal retinal elevation, subretinal fluid, focal vitreous traction over the temporal retina (closed arrowhead) and subretinal band (open arrowhead). The location of the cross-sectional OCT scan was marked with green line on the en face scan inset.

**Figure 4.**
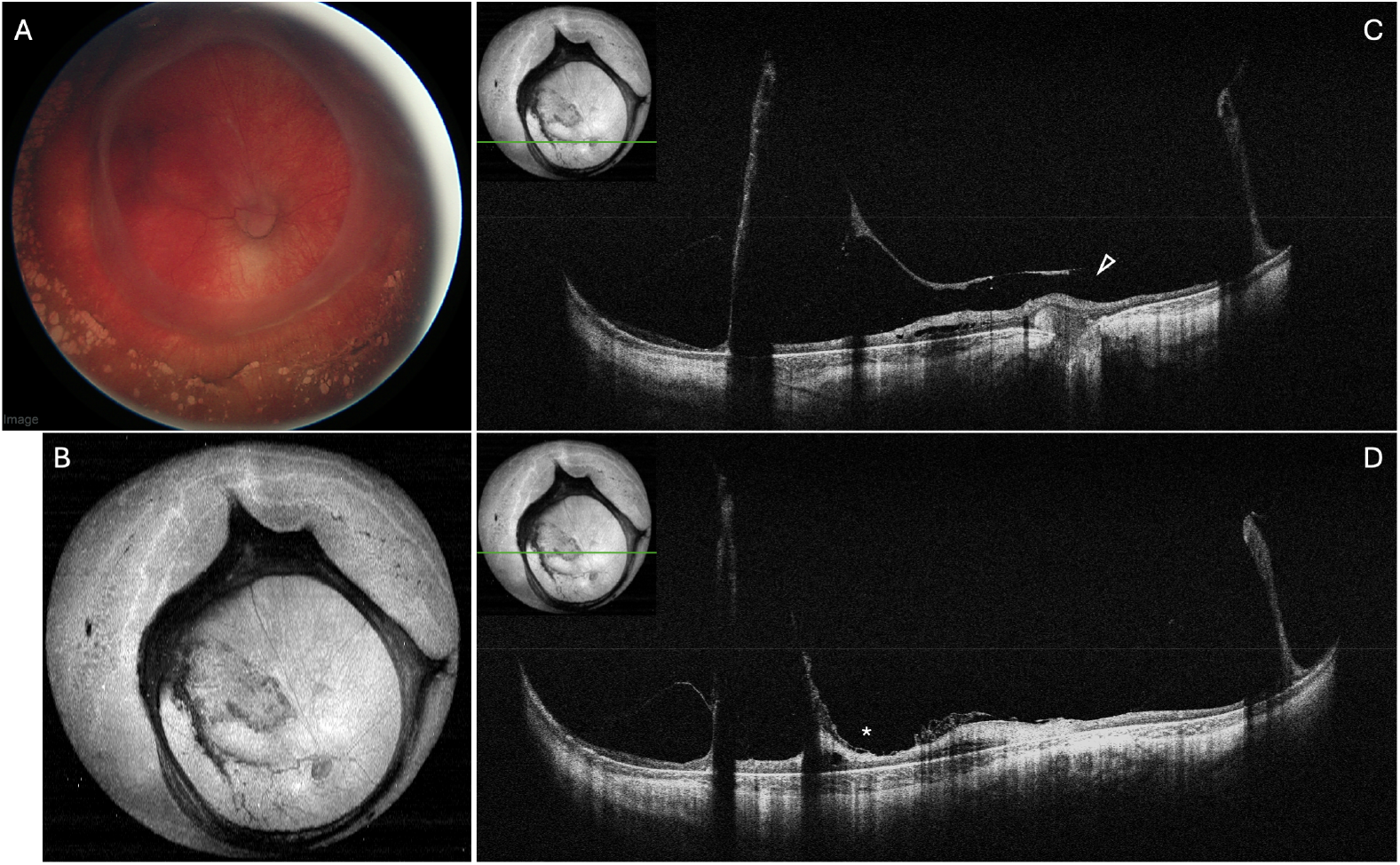
Right eye of a preterm infant imaged under anesthesia following surgical repair of stage 4B retinopathy of prematurity. Widefield fundus photograph (A) and widefield en face OCT (B) showed ring-like contraction of the vitreous and retina at the vascular-avascular junction. Cross-sectional OCT scan through the optic nerve (C) and the macula (D) showed attached retina with residual vitreous traction, optic nerve elevation (open arrowhead) and tractional near the central macula (asterisk). The location of the cross-sectional OCT scan was marked with green line on the en face scan inset.

**Figure 5.**
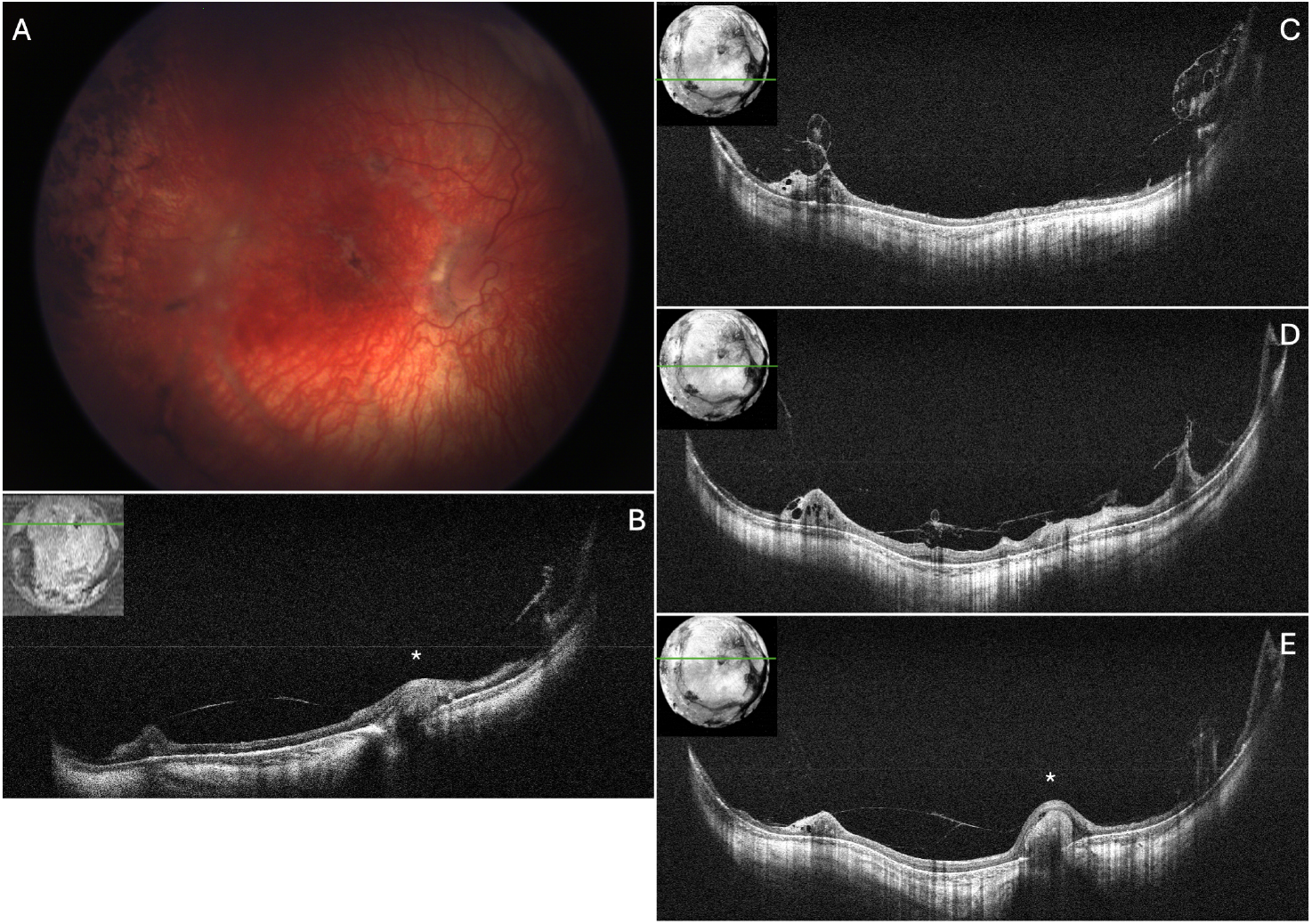
Right eye of a preterm infant following surgical repair of stage 5 retinopathy of prematurity imaged in clinic and under anesthesia 5 months after the clinic visit. Widefield fundus photograph (A) showed attached posterior pole with residual vitreous traction. Cross-sectional OCT scan obtained in clinic (B) and under anesthesia 5 months after the clinic visit (C-E) showed attached retina with residual, multifocal vitreous traction, optic nerve elevation (asterisks) and tractional retinoschisis and multifocal retinal thickening. The vitreous traction over the macula appeared relatively unchanged over the 5 months interval (comparing B and E). The location of the cross-sectional OCT scan was marked with green line on the en face scan inset.

**Figure 6.**
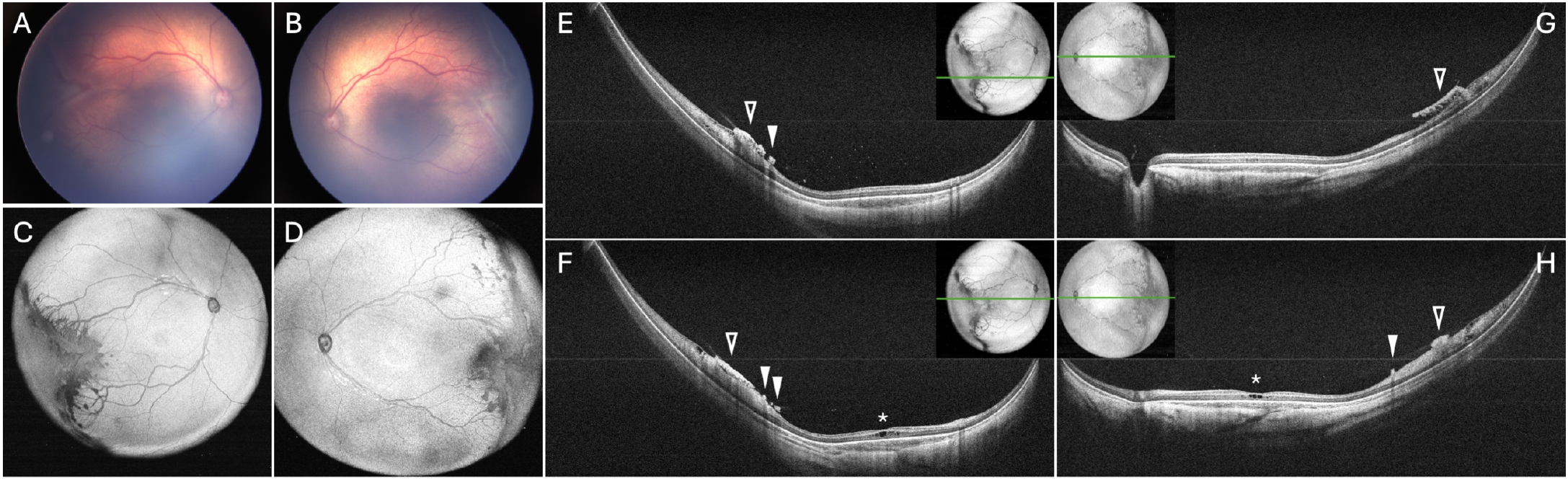
Both eyes of a preterm infant with stage 3 retinopathy of prematurity imaged at the bedside in the nursery. Widefield fundus photograph (A,B) and widefield en face OCT (C,D) showed temporal elevated neovascular ridge at the vascular-avascular junction. Cross-sectional OCT scan through the macula (E,G) and the fovea (F,H) showed vitreous hyperreflective foci, temporal retinal thickening and neovascular elevation with associated vitreous traction, cystoid macular edema (asterisks, F, H), neovascular buds (closed arrowheads) and neovascular plaque (open arrowheads). The location of the cross-sectional OCT scan was marked with green line on the en face scan inset.

### Widefield cross-sectional visualization of the retinal features

Intraretinal features visualized on widefield cross-sectional OCT scans include retinal break with localized subretinal fluid in a child with an operculated hole anterior to the equator **(Figure 1H)**, intraretinal hyperreflective foci and retinal thickening in a child with Coats’ disease **(Figure 2D-E)**, retinal thickening near the vascular-avascular junction in a preterm infant with stage 3 ROP **(Figure 2D and Figure 6E-H, Supplementary video 1)**, and tractional retinoschisis in multiple subjects **(Figure 3C, Figure 4C,D, Figure 5B-E and Figure 6E-H)**.

### Widefield cross-sectional visualization of subretinal features

Subretinal features visualized on widefield cross-sectional OCT scans include subretinal exudates **(Figure 2D, E)** and subfoveal nodule in a child with Coats’ disease **(Figure 2E)**, subretinal band in chronic retinal detachment **(Figure 3D)**, and optic nerve elevation in stage 5 ROP status post retinal detachment repair **(Figure 5E)**. Choroidal features are well-visualized in infants with swept-source OCT, especially at the posterior pole, but visualization of both retinal and choroidal features appear to taper off toward the edge of the cross-secPonal OCT scan.

## Discussion

In our pilot study, application of the investigational handheld OCT system with widefield lens allowed high-quality visualization of both en face and cross-sectional OCT information, providing a three-dimensional evaluation of the vitreoretinal interface. It provides us with important structural relationship between the vitreous and the retina not only at the posterior pole but also at the retinal periphery. At the same time, retinal and subretinal features are also visualized with high quality.

With the advent of the spectral-domain handheld OCT system and the recent FDA approval of the Heidelberg FLEX system for supine OCT imaging, physicians have a larger repertoire to evaluate the pediatric retina. However, both systems are limited by the FOV and time of capture, making it difficult to capture beyond the posterior pole, or capture OCT volume information at the bedside. Several investigational swept-source OCT systems have shown promises for faster and widefield evaluation of the pediatric retina,^13,15,18^ as well as capability of OCT angiography.^14,16-19^ Here we report and evaluate the use of a pre-commercial handheld OCT system with widefield lens, which show promises in evaluation of the pediatric retina with both en face and cross-sectional OCT information.

There are limitations of this and other handheld OCT systems. Compared to fundus flash photography, the capture of OCT volumes, despite at a faster speed, still requires 1-2 seconds per volume, which in awake infants, could lead to some motion artifacts. This is an issue for any OCT system and can be overcome by comparing repeated volumes. Toward the edge of the OCT volumes, visualization of both retinal and choroidal features could be tapered off and thus may limit interpretation of retinal and choroidal thicknesses at the edge of the OCT volume.

In summary, the use of this new pre-commercial investigational handheld OCT system with widefield lens provide valuable en face and cross-sectional information of the pediatric retina. A wider application of this system may improve our understanding of the development and pathology of pediatric retinal diseases.

## Methods

### Subjects

This pilot study was approved by the Duke Health Institutional Review Board and adheres to the Health Insurance Portability and Accountability Act and all tenets of the Declaration of Helsinki. Pediatric subjects undergoing clinical examination or clinically indicated examination under anesthesia at Duke University Hospital or Duke Health Clinic were enrolled with written informed consent from parents/legal guardians.

### Widefield OCT imaging

The Theia T1-W investigational handheld OCT system utilizes a 1060 nm laser and has a scanning speed up to 500 frames/second with a FOV of 145 degrees, allowing for the acquisition of full retinal volumes in infants with a large field of view. The image acquisition time was approximately 1.2 to 2.4 seconds per OCT volume. The system consists of a cart-mounted swept-source engine connected to a lightweight (350 gram) ergonomically designed handheld probe using a flexible tether. During imaging, a reusable contact lens is placed on the eye with a coupling gel which aids in stabilization of the eye. The contact lens was sterilized between imaging sessions. All images were acquired by trained photographers at the bedside or during examination under anesthesia. OCT volumes were optimized for either high resolution cross-sectional B-scan imaging or en face image quality. The volumes were processed with custom MATLAB software. The FOV was visually evaluated by comparing the en face OCT images to the standard-of-care widefield (RetCam) or ultra-widefield (Optos) retinal fundus images.

## Data Availability

All data produced in the present study are available upon reasonable request to the authors.

